# Sex and menstrual cycle differences in the mood-activity association should inform cycle-aware digital phenotyping

**DOI:** 10.64898/2026.07.24.26358675

**Authors:** Kyra Delray, Jakob K Zeitler, Joseph F Hayes, Aaron Kandola, Nicky Keay, Robin J Evans

## Abstract

Passive physical activity is increasingly used as a digital-phenotyping proxy for mood, assuming a stable relationship across people and time. We tested this assumption using daily mood and activity data from 1,072 individuals (121 women, 951 men; 13,909 person-days) in the Juli app.

The within-person activity-mood association was stronger in women than men (slope difference −0.194, *p* = 0.019). Within women, it varied across the menstrual cycle (*p* = 0.022): absent in the early luteal phase, significant in all other phases, and largest in the late luteal phase (+0.46). A male pseudo-cycle control showed no such modulation (*p* = 0.974), confirming the effect is cycle-specific rather than a general temporal pattern.

Accounting for cycle phase improved out-of-sample mood prediction in women in 75% of cross-validation splits. Digital phenotyping should account for sex and menstrual cycle phase to avoid biased predictions and enable personalized recommendations for women.

## Introduction

Digital health technologies (DHTs) make it possible to continuously monitor a person’s state over long periods in their natural environment, rather than relying on one-off measurements in a clinical setting. This offers a more granular view of a patient’s condition, and when combined with environmental context such as GPS and self-report surveys, allows researchers to understand how a person’s environment affects their symptoms and severity. In turn, this can help researchers design real-world behavioural or environmental interventions.

Digital phenotyping [1, 2] is a recently introduced concept describing the quantification of information captured by DHTs to infer a person’s state. It encompasses the real time quantification of behaviour, cognition, and mood from data passively collected by personal devices such as smartphones and wearables.

Previous work has shown that ecological momentary assessments (EMAs) of mood can statistically represent variation in clinically validated depression questionnaires [3]. This means a brief, regularly repeated survey can serve as a good proxy for depressive symptoms: a powerful tool for understanding how changes in a person’s behaviour or environment relate to changes in their depressive symptoms. The ability to discover passive markers correlated with this EMA signal is therefore especially appealing, since such markers would place an extremely low burden on the user. Passive behavioural monitoring has, as a result, become a central tool in digital mental health research: smartphones and wearables continuously record physical activity, sleep, location, and social behaviour, and these signals are increasingly used to monitor mood and mental health symptoms without requiring active self-report [4].

Physical activity is among the most widely used passive proxies for mood. A large meta-analytic literature establishes a positive overall association between physical activity and mood or depressive symptoms [5–7]. At the individual level, within-person increases in activity tend to track with better same-day or next-day mood [3]. These findings have motivated the use of passively recorded step counts and activity data as real-time mood indicators in digital health platforms.

To act on the information provided by these digital phenotypes, such signals can be formalised within Dynamic Treatment Regimes (DTRs) [8], a general framework for varying treatment decisions across time. Just-in-time adaptive interventions (JITAIs) are a specific application of this idea: they aim to deliver the right type and amount of support at the right time by adapting to a person’s changing internal and contextual state, using passively sensed data and self-report [9].

The assumption underlying this use is that the activity-mood association is approximately stable: a given increase in activity corresponds to a constant improvement in mood across time, across people, and within a person across different physiological states. If that relationship varies systematically between people, or in women across the menstrual cycle, a fixed decision rule will be biased and its recommendations would be systematically suboptimal.

There is evidence that the association varies by sex [10], with some studies reporting a stronger link in women, and evidence that mood itself varies across the menstrual cycle [11–13]. In our own prior work using ecological momentary assessment in this population, mood changed systematically across the menstrual cycle [3]. Hormonal fluctuations across the cycle are well documented and thought to underlie this mood variation [12].

What has not been examined is whether the activity-mood association varies within women across the cycle. If it does, a model that treats the association as constant will be systematically incorrect across the cycle.

We address three questions. First, does the within-person same-day activity-mood association differ between women and men? Second, within women, does this association vary across menstrual cycle phase? Third, does accounting for the cycle improve out-of-sample prediction of mood from activity? We use a within-person fixed-effects design, a male pseudo-cycle negative control to distinguish cycle-specific from general temporal effects, a Fourier-basis varying-coefficient model to characterise the continuous shape of the association across the cycle and support interpretable, day-level cycle-adjusted phenotyping, and cross-validation to test predictive power.

## Results

### 1.1 Sample

The sex-comparison model comprised 1,072 individuals (121 women, 951 men; 13,909 person-days). The cycle-modulation analysis used 121 women with at least two documented menstrual cycles and same-day mood and activity data. These 121 women contributed 3,512 analysis days (a median of 8 days per woman, interquartile range (IQR) 3-18) across a median of four recorded period onsets. 76% of onsets were algorithmically imputed by extrapolating from cycle length between recorded menstruation onsets.

The cross-validation analysis used 121 women (3,512 person-days) and 951 men (10,397 person-days). Figure 1 shows the distributions of mood and activity before and after within-person normalization.

**Figure 1:**
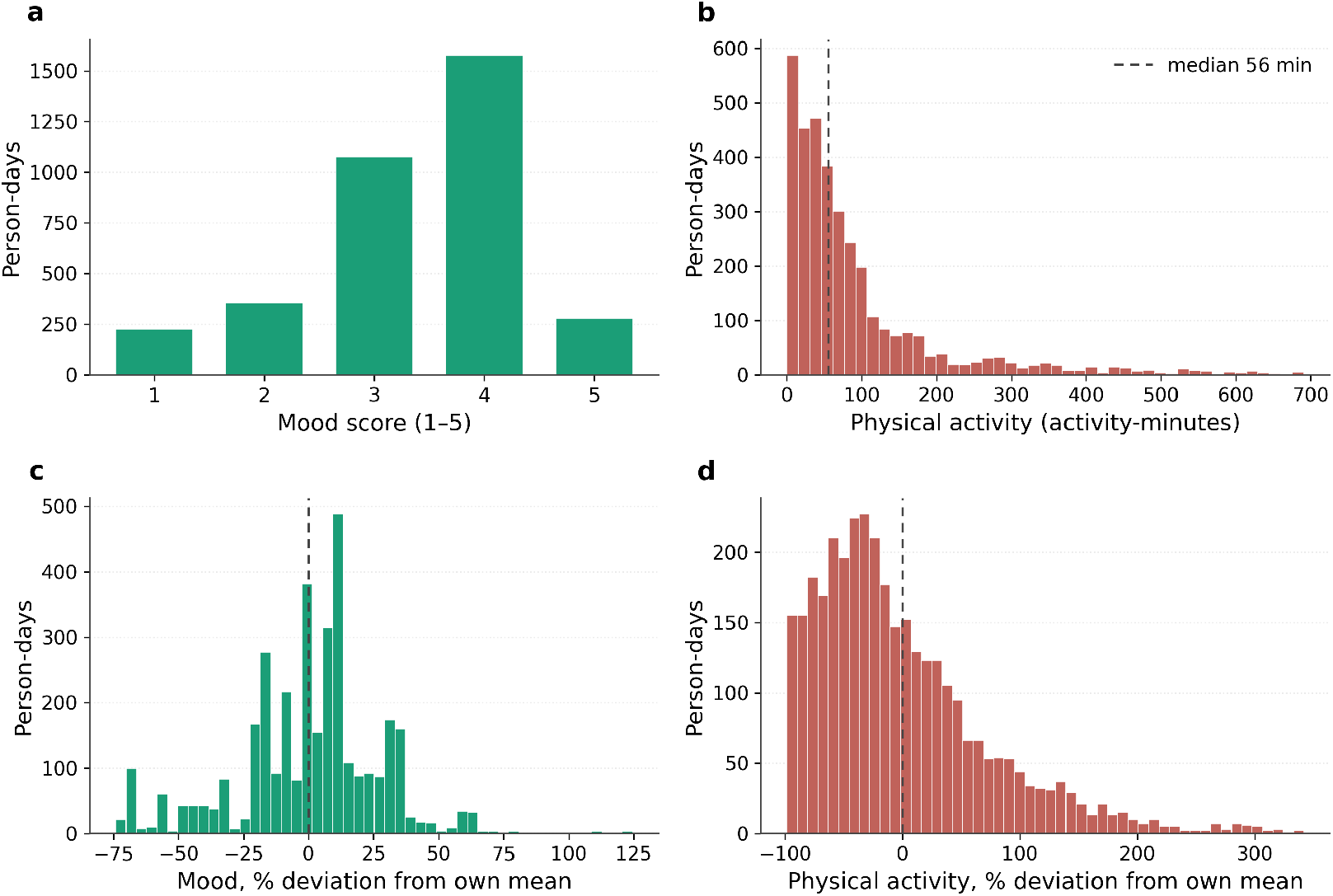
Distributions of daily mood and physical activity before and after within-person normalization. Analytic sample of 121 women, 3,512 person-days. (**a**) Raw mood on a five-point scale (1 = very bad to 5 = excellent), teal bars. (**b**) Raw physical activity in activity-minutes (step count divided by 100 plus workout minutes), terracotta bars; the dashed line marks the median (56 activity-minutes). (**c, d**) The same two variables as within-person percentage deviations from each woman’s own mean, the form entered into all models; dashed lines at zero. Teal denotes mood and terracotta physical activity throughout; vertical axes show person-days. Activity remains right-skewed after normalization.

### 1.2 Overall activity-mood association and sex difference

Pooled across all participants, the within-person same-day activity-mood association was positive but small. In the pooled model (*n* = 1,072; 13,909 person-days), the slope was +0.289 in women and +0.095 in men. The difference (men minus women) was −0.194 (95% CI [−0.356, −0.032], *p* = 0.019), indicating the association was reliably stronger in women.

### 1.3 Cycle modulation within women

Within the female subsample (*n* = 121), the activity-mood association varied across the menstrual cycle. The omnibus phase × activity interaction was statistically significant (*p* = 0.022). Per-phase estimates are given in Table 1 and shown in Figure 2.

**Table 1:** Within-person physical activity-mood slope by menstrual cycle phase (*n* = 121 women). Person fixed-effects OLS with mood × cycle-phase interaction. Omnibus interaction *p* = 0.022.

| Phase (cycle days) | Slope | 95% CI | CI excludes 0 |
| --- | --- | --- | --- |
| Early luteal ($-14$ to $-9$) | 0.00 | $[-0.31, +0.30]$ | No |
| Late luteal ($-8$ to $-1$) | +0.46 | $[+0.21, +0.71]$ | Yes |
| Menstruation (0 to 5) | +0.36 | $[+0.15, +0.57]$ | Yes |
| Follicular (6 to 13) | +0.27 | $[+0.12, +0.43]$ | Yes |

**Figure 2:**
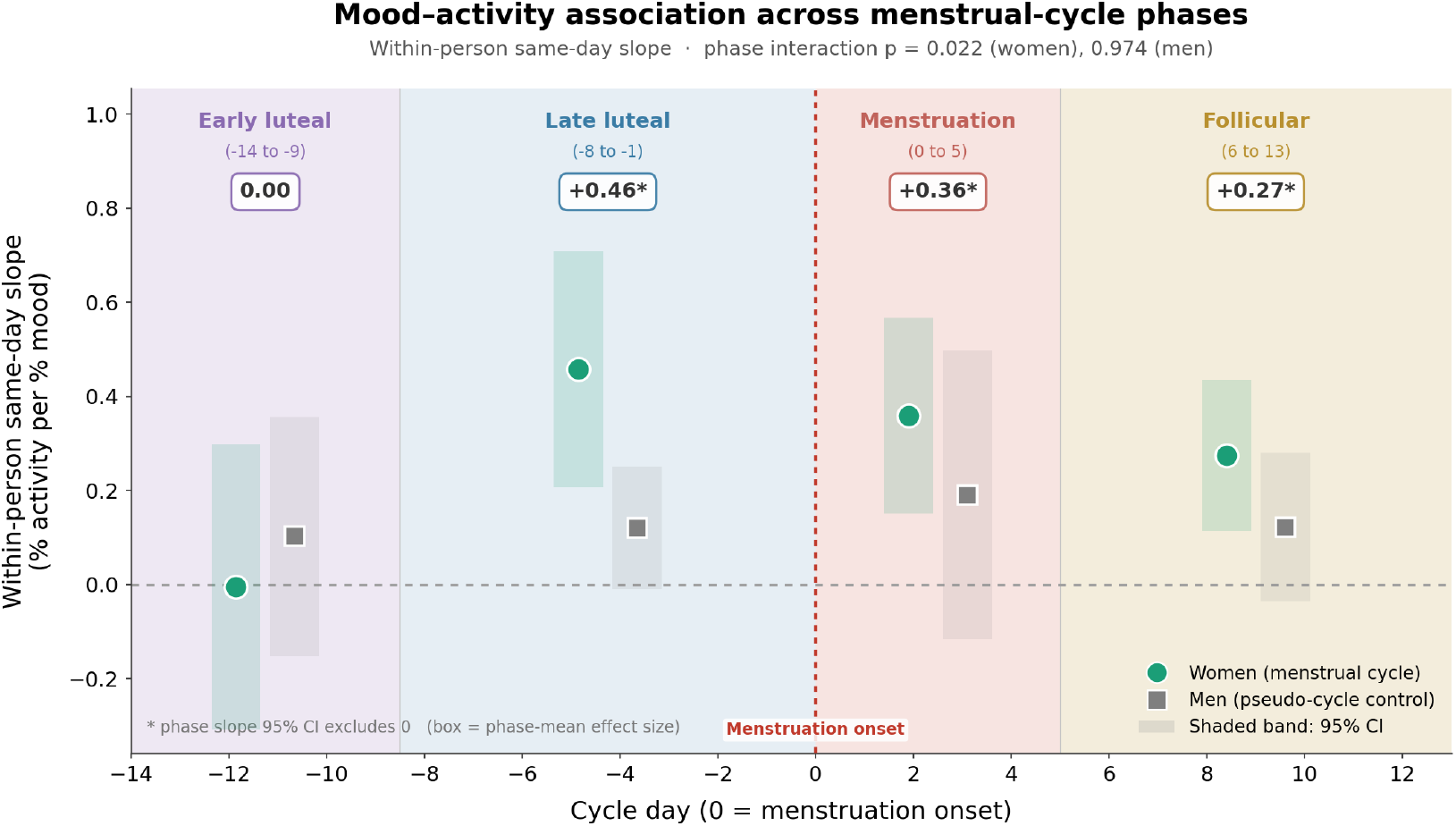
Within-person physical activity-mood slope by menstrual cycle phase, with a male pseudo-cycle negative control. Each marker is the within-person, same-day slope relating physical activity to mood, estimated by person fixed-effects ordinary least squares with standard errors clustered by individual and adjustment for day of week and daily weather (temperature, humidity, atmospheric pressure, pollen). The slope is the percentage change in activity per one-percent change in mood, so positive values indicate that a woman’s above-average mood days are also her above-average activity days. Teal circles denote women (*n* = 121) at their observed menstrual cycle phase; grey squares denote men (*n* = 707) at synthetic pseudo-cycle phases of matched width, anchored to the first Monday of each calendar month. Shaded bands around each marker show 95% confidence intervals; they are not standard errors or standard deviations. Each plotted slope is a single estimate representing the whole phase. The horizontal axis is cycle day, with day 0 the first day of menstruation, marked by the red dotted vertical line; the grey dashed horizontal line marks a slope of zero. Background shading indicates the four cycle phases (purple, early luteal; blue, late luteal; red, menstruation; yellow, follicular). The boxed value above each phase is the phase-mean slope for women, marked with an asterisk where its 95% confidence interval excludes zero. In women the slope is near zero in the early luteal phase and positive in the late luteal, menstruation and follicular phases; in men it is flat across all four pseudo-phases. A sample-size-matched version of the male control is reported in the sensitivity analyses.

The association was zero in the early luteal phase and significant in all other phases, with the largest point estimate in the late luteal window. The follicular and menstruation confidence intervals overlap substantially; we do not claim ordering among them beyond what the formal omnibus test supports.

### 1.4 Continuous view across the cycle

The Fourier-basis varying-coefficient smooth (Figure 3) provides a bin-free view of the same pattern. The slope rises from near zero in the early luteal window, peaks around the late luteal phase and menstruation, and remains elevated through the follicular phase before declining. The grey dashed line for men is flat across the same window. Note that the pointwise confidence bands for the two continuous smooths are not directly compared to each other, so their overlap here should not be read as evidence against a real difference; the formal test for that difference is the omnibus phase × activity interaction on the binned model (Table 1), which is significant in women and null in the male pseudo-cycle control. Given the female sample is far smaller than the male one, we would expect this overlap to narrow with a larger female sample. The boxed values in Figure 3 come from the binned phase model; the curve is the Fourier smooth. These are related but distinct estimators of the same underlying variation.

**Figure 3:**
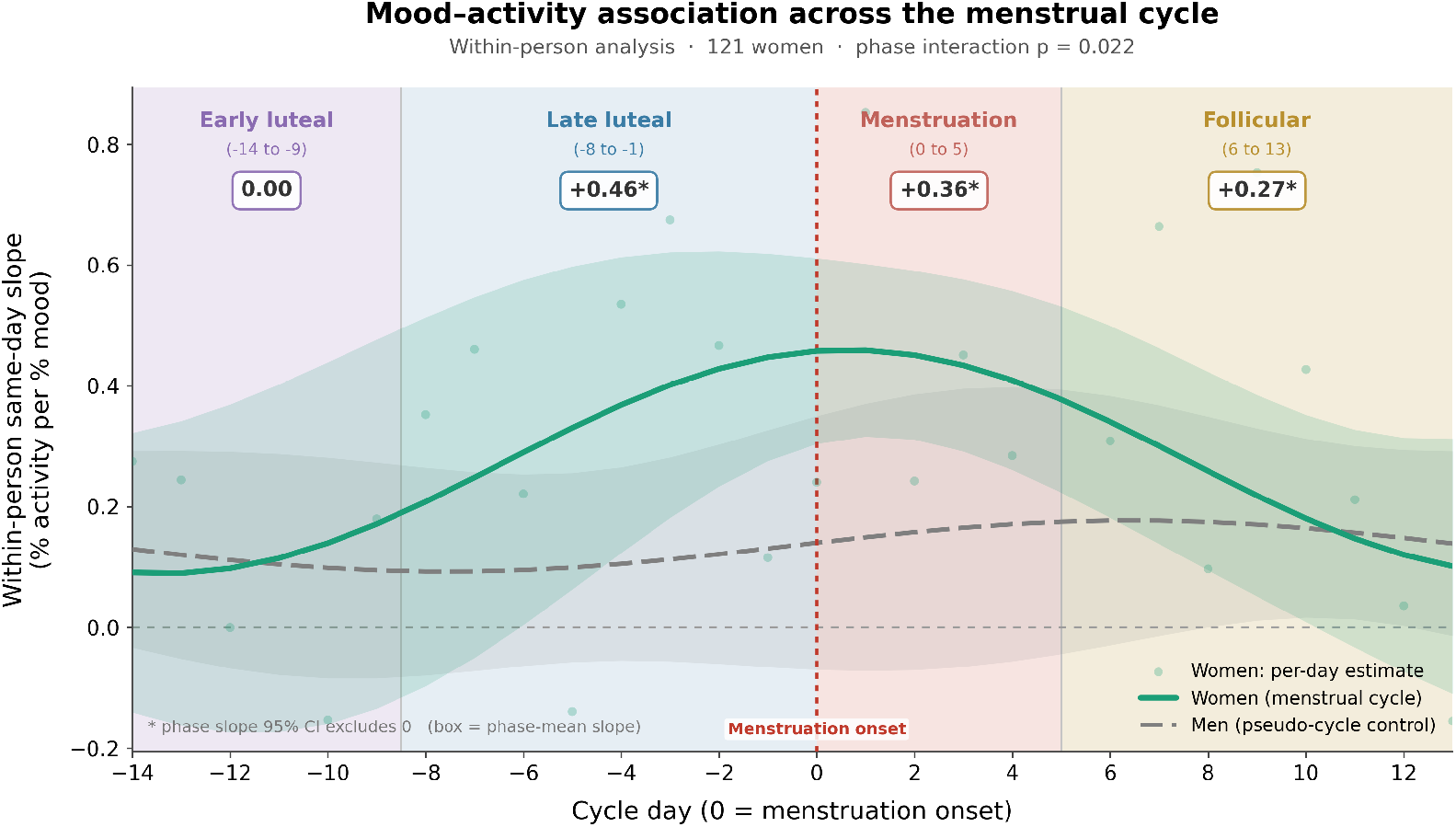
Continuous variation in the within-person physical activity-mood slope across the menstrual cycle. The same-day within-person slope relating physical activity to mood, modelled as a smooth periodic function of cycle day rather than in discrete phases. As in Fig. 2, the slope is the percentage change in activity per one-percent change in mood, estimated with person fixed effects, standard errors clustered by individual, and adjustment for day of week and daily weather. The phase-specific slope was then modelled as a smooth function of cycle day using a generalized additive model (GAM) with a Fourier basis. The solid teal line is the fitted slope for women (*n* = 121; 3,512 person-days), surrounded by a teal shaded band showing the pointwise 95% confidence interval. The dashed grey line is the corresponding fit for men (*n* = 707; 8,108 person-days), with a grey shaded band showing its pointwise 95% confidence interval; men were assigned synthetic pseudo-cycles anchored to the first Monday of each calendar month. Faint teal dots are the slope estimated independently for each of the 28 cycle days, without smoothing, from the same model and covariates, plotted for women only. The horizontal axis is cycle day, with day 0 the first day of menstruation, marked by the red dotted vertical line; the grey dashed horizontal line marks a slope of zero. Background shading indicates the four cycle phases (purple, early luteal; blue, late luteal; red, menstruation; yellow, follicular). Boxed values are the phase-mean slopes from the binned model, the same estimates shown in Fig. 2 and Table 1, asterisked where the 95% confidence interval excludes zero.

### 1.5 Negative control: male pseudo-cycle

In male users, the overall within-person activity-mood association was positive and significant (+0.137, *p* = 0.017; this is slightly larger than the male slope in the pooled sex comparison in Section 1.2 because the negative-control model additionally adjusts for weather and is restricted to weather-complete records). However, when the identical phase-modulation pipeline was applied using synthetic 28-day pseudo-cycles, there was no variation across pseudo-phases (*F* (3, 706) = 0.07, *p* = 0.974). This evidence suggests that the modulation observed in women is a result of the menstrual cycle rather than a general temporal effect.

### 1.6 Cycle-aware modelling and out-of-sample mood prediction

Finally, we considered whether accounting for the cycle actually improves the out-of-sample prediction of mood from activity for future use in digital phenotyping and JITAIs. In repeated five-fold cross-validation, allowing the activity coefficient to vary by cycle phase increased held-out *R*^2^ in women in 75% of the cross-validation splits (mean gain +0.054 percentage points). In the male pseudo-cycle control did not improve prediction accuracy (positive in 0% of splits; mean −0.060 percentage points). We also note that the predictive increase does not depend on the direction of prediction: reversing the model to predict activity from mood also increased predictive accuracy, a gain in women in 80% of splits and none in men (0%). Cycle-aware modelling therefore consistently recovers predictive mood signal in women compared to a non-cycle-aware model.

### 1.7 Sensitivity analyses

We examined whether the cycle-modulation result depended on modelling choices. Cycle day was assigned from period start dates, of which 76% in the analysed cohort were algorithmically imputed rather than user-reported through cycle length extrapolation. Restricting the cycle-day labelling to observed onsets only reduced the sample to 111 women and 2,500 person-days but preserved the pattern: the phase × activity interaction remained (*p* = 0.047), the early luteal slope stayed near zero, and the late luteal, menstruation, and follicular slopes remained positive. The modulation is therefore not a result of the imputation.

The result was robust to how physical activity and mood were quantified. Varying the step-to-minute cadence used to combine steps with workout minutes left the phase interaction essentially unchanged (*p* between 0.013 and 0.05), and it was strongest when steps were used alone (*p* = 0.005).

Removing the weather covariates from the model, which therefore relaxes the requirement that weather data be collected on the same day as the observation, retains 134 women (3,976 person-days). This strengthened the significance of the omnibus phase interaction to *p* = 0.006 and the per-phase slopes were similar (early luteal −0.05; late luteal +0.39; menstruation +0.38; follicular +0.29). Because the coefficients are stable, the stronger significance is most plausibly attributable to the larger sample. We present weather in the primary model to ensure we account for any possible confounding.

We also asked whether the pattern depended on how much each woman logged or how active she was, splitting the sample at the median of each. Among women who logged more (61 women, 3,301 person-days), the phase modulation matched the full-sample result (*p* = 0.013). However, among those who logged few observations (60 women, 211 person-days, roughly 3.5 usable days each), no association was estimable; we believe this reflects insufficient statistical power from the small sample.

Split instead by activity level, the cycle modulation was concentrated in less active women (*p <* 0.001) and weaker in habitually active women (*p* = 0.21), despite the latter contributing more than twice as many person-days. More frequent recorders were also more active (Spearman *r* = 0.29, *p* = 0.001), so the two splits are correlated; these subgroup analyses are exploratory, and formally testing whether activity level moderates the cycle effect would require a three-way interaction with a larger sample size for meaningful inference.

The male pseudo-cycle control is estimated on a larger sample than the female analysis (707 versus 121); the male count reflects the same minimum per-phase data criteria used throughout, which reduced it from the full cohort of 951. Because males do not have to have self-reported a menstrual cycle, the sample size is naturally larger. For completeness, we re-estimated the male per-phase slopes on random subsamples matched to the female sample size (*n* = 121, averaged over five draws). At matched size the male confidence intervals widen to include zero at every phase, and the overall male slope is no longer distinguishable from zero (+0.13, 95% CI [−0.18, +0.43]); the phase interaction remains null (*p* ≈ 0.40). The absence of phase modulation in men is therefore not related to sample size.

## Discussion

This work provides evidence that digital phenotyping of physical activity for mood is biased if it does not appropriately account for sex and the menstrual cycle. The effect is stronger in women than men, and within women it varies across the menstrual cycle: there is no relationship between mood and PA in the early luteal phase, but a statistically significant association in all other phases. The male pseudo-cycle control shows that this effect is related to the menstrual cycle itself, and not simply to the time of month. Allowing the association to vary across the menstrual cycle also increased predictive accuracy in women relative to a non-cycle-aware model.

For digital phenotyping systems, this implies that cycle phase should be incorporated as a moderator when physical activity is used as a mood signal in women. As just-in-time adaptive intervention systems become more sophisticated, cycle-aware models offer a way to improve the quality of the behavioural phenotyping.

The finding that the activity-mood association is stronger in women than men (slope difference −0.194, *p* = 0.019) is consistent with prior evidence of sex heterogeneity in the physical activity-mental health literature [10]. Further work is necessary to understand whether this is biological, environmental, or sociological in origin.

Menstruation onset (cycle day 0) is our anchoring event, directly reported by the user, and previous work estimates menstruation to last an average of five days [14]. The follicular phase is the most variable window in our analysis, spanning the days from +5 until 14 days before the next cycle’s onset (a span of between 2 and 14 days depending on the user’s cycle length). The luteal phase begins 14 days before the next cycle’s onset, based on research showing that ovulation generally occurs at this point regardless of overall cycle length [15]. However, the boundary between the early and late luteal phase is not attached to a specific physiological event, so we placed it where the coupling changed rather than fixing it in advance. This boundary is therefore best read as a data-driven estimate of where the coupling begins to strengthen.

This phase corresponds to what we expect to be the immediate post-ovulatory period, when progesterone rises sharply [15]. The mechanisms by which hormonal variation might modulate the activity-mood coupling are speculative and not established by these data, and we do not assert a hormonal explanation.

One possible explanation, offered here as a hypothesis rather than a finding confirmed by our data, draws on established endocrinology of the menstrual cycle. The luteal phase of the menstrual cycle is the time of the most dynamic changes in the ovarian sex steroids estradiol and progesterone. These hormones are not restricted to physical effects, as they are able to cross the blood-brain barrier and interact with neurotransmitters, impacting mood [16]. This flux of ovarian hormones in the luteal phase corresponds to the timing of pre-menstrual syndrome (PMS). PMS does not occur where there is no fluctuation in ovarian hormones [17]. Each woman will have her own biological response to hormone fluctuations, which is why PMS is a clinical diagnosis based on symptoms, not hormone blood tests. Some women appear particularly sensitive to hormone fluxes occurring in PMS, post-natal depression and perimenopause, and are subsequently susceptible to depression in these parts of the female hormone odyssey [18].

These individual differences in hormone sensitivity have direct implications for how digital health tools should be designed. As women have individual sensitivities to ovarian hormones, and PMS and perimenopause are diagnoses based on symptoms rather than hormone levels, digital phenotyping systems cannot rely on a uniform threshold to flag or personalize mood-related recommendations for women: the same passive signal may call for a different response depending on where a woman is in her cycle. Rather than treating physical activity as a one-size-fits-all intervention, digital tools should personalize recommendations, accounting for factors such as baseline fitness, individual exercise preferences, and cycle phase.

The sample is self-selected: Juli users who actively track mood and connect activity and cycle data are not representative of the general population or of clinical populations. Mood is a single-item daily self-report; while validated against the PHQ-8 in this dataset [3], it does not capture the full dimensionality of mental health. Cycle phase is derived from self-reported period start dates with no hormonal confirmation and no ovulation marker [12]; the luteal phase and follicular phase boundaries are estimated, not measured. Physical activity is device-measured and subject to wear compliance and device heterogeneity. The phase-modulation analysis is based on *n* = 121 women and replication in larger, more diverse cohorts is warranted. The out-of-sample predictive gains, while consistent in direction, are small in absolute terms (under 0.1 percentage points of held-out *R*^2^), so the practical value of cycle-aware modelling should be confirmed prospectively.

Sleep data were available for only approximately 2% of cohort-days. An exploratory analysis of the sleep-mood association across the cycle was uninformative: the overall sleep-mood slope was near zero (−0.01, *p* = 0.84) in only 20 women with usable data. As sleep is also related to the menstrual cycle [19], this should be considered in future analysis where more sleep data is available.

Digital phenotyping should make efforts to incorporate menstrual cycle data for women, for unbiased and equitable dynamic treatment regimes, JITAIs, and personalized recommendations. Where menstrual cycle data is not available, it should at least consider the sex of an individual, as the association appears to be stronger in women than men. A male pseudo-cycle negative control confirms the modulation is menstrual-cycle-specific, and cycle-aware modelling improved predictive accuracy and recovered out-of-sample mood signal in women that a non-cycle-aware model discards. Though here we tested only physical activity and mood, it is possible this pattern may exist in other digital phenotypes. Digital health apps should therefore consider implementing cycle-aware phenotyping for better accuracy and more useful recommendations for women.

## Methods

### 2.1 Data source

We analysed data collected from Juli, a chronic health management digital application available on iOS and Android. Users voluntarily log daily mood ratings and may connect their accounts to Apple HealthKit or Google Fit for physical activity data. Menstrual cycle data (period start dates) are imported directly from Apple Health or Google Health Connect.

The data were provided by Juli Inc; one author (J.F.H.) is a co-founder of Juli Inc.

#### Ethics approval

This study uses data from the same Juli cohort as our related analysis [3] and is covered by the same ethical approval: University College London Research Ethics Committee (ID 19413/002). Users provided informed consent for their deidentified data to be used for research during the Juli app onboarding process.

### 2.2 Measures

#### Mood

Daily mood was rated on a five-point scale (1 = very bad, 5 = excellent). Scores were normalised to each individual’s own mean across all observations, expressed as percentage deviation, so that person-level means are zero by construction and within-person fluctuations are the unit of analysis. This measure has been shown to be significantly associated with the PHQ-8, the typical clinical questionnaire for depression, in this dataset [3].

##### Physical activity

Daily activity was obtained passively from smartphone sensors or linked wearables via HealthKit or Google Fit, primarily as step counts, supplemented by self-recorded exercise sessions [20]. We divided daily step count by 100 to approximate minutes of walking, based on a cadence of roughly 100 steps per minute for moderate-intensity walking [21], and added minutes from recorded workout sessions. Physical activity minutes were normalised to each person’s own mean, expressed as percentage deviation.

##### Menstrual cycle phase

Preprocessing and cyclic modelling followed the conceptual framework of the open-source mcanalysis package [22]. Women were included if they had a cycle of normal length (21-35 days), which requires at least two recorded period onsets, and contributed at least one day on which mood, physical activity and weather were all recorded. Of 1,013 women in the cohort, 161 met the cycle-length criterion and 121 additionally contributed at least one complete day, forming the analytic sample. Period start dates were used to define cycle day 0 as the first day of menstruation, with preceding days assigned negative values. The analysis window was −14 to +13. Four phases were defined based on established hormonal landmarks [12] or prior work [3]: early luteal (days −14 to −9), late luteal (−8 to −1), menstruation (0 to 5), and follicular (6 to 13).

### 2.3 Statistical models

#### Primary model

Our primary analysis tested the same-day relationship between physical activity (PA) and mood. This relationship was estimated as the slope of a within-person fixed-effects ordinary least squares (OLS) regression, with standard errors clustered by person. Normalised physical activity was regressed on normalised mood, each expressed as a percentage deviation from the person’s own mean, so the coefficient of interest is the same-day within-person slope: the percentage change in activity associated with a one-percent change in mood (the quantity plotted in Figures 2 and 3).

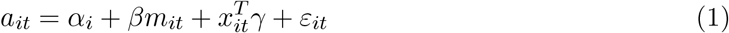

The model adjusted for day-of-week indicators and daily weather (temperature, humidity, atmospheric pressure, and pollen). Because activity and mood are recorded on the same day and centered within person, the estimate is a contemporaneous within-person association and is not interpreted as a causal effect.

##### Sex comparison

A pooled model including all participants was fitted with a sex × physical activity interaction term (*n* = 1,072 people; 13,909 person-days).

##### Cycle-phase interaction model

Within the female subsample (*n* = 121), a mood × cycle-phase interaction was added. The omnibus test used a cluster-robust Wald F-statistic to test whether the slope differed significantly across phases.

##### Continuous model

To describe the association across the cycle without splitting it into discrete phases, and to allow adjustments at the daily level, we fit a smooth across the whole cycle, estimating the same-day activity-mood slope at each cycle day. The smooth is fit on a canonical 28-day cycle, the mean cycle length; because it is periodic, the fitted curve can then be stretched or compressed to match an individual’s own cycle length when the model is applied, for example expanded across a 30-day cycle. For person *i* on day *t*, with *a*_*it*_ and *m*_*it*_ the within-person percentage deviations in activity and mood,

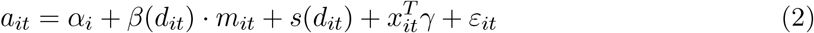

where *d*_*it*_ is cycle day, *α*_*i*_ is a person fixed effect, and *x*_*it*_ holds the day-of-week and weather covariates. The slope *β*(*d*) and the activity-level term *s*(*d*) are each modeled as a Fourier series in cycle day: a sum of sine and cosine terms with a 28-day period. Because sine and cosine are periodic, the fitted curves start and end at the same point and so are continuous around the cycle, wrapping around smoothly instead of jumping at the cycle boundary. This lets the model be evaluated at any day of an ongoing cycle, not just within fixed phase bins, so the cycle can be treated as a continuous trend and applied appropriately in real-world settings.

The number of harmonics was tested among *K* = 1, 2, 3 and selected using the Bayesian Information Criterion to avoid overfitting. The slope *β*(*d*), plotted in Figure 3, shows how the activity-mood association changes across the cycle; standard errors clustered by person give its pointwise 95% bands.

##### Negative control

The identical pipeline was applied to male users, using the first Monday of each calendar month as the pseudo-cycle onset. The aim was to test whether a cyclical temporal effect exists independent of the menstrual cycle.

##### Prediction utility check

To assess whether adjusting digital phenotyping for the menstrual cycle is useful in practice, we tested whether accounting for cycle phase improves out-of-sample inference of mood from activity. Within person, we predicted mood from activity and compared a non-cycle-aware model with one in which the activity coefficient could vary by cycle phase, assessing performance via held-out *R*^2^ under repeated five-fold cross-validation (40 repetitions). We report the change in accuracy from letting the coefficient vary by phase, and ran the identical comparison in the male pseudo-cycle cohort as a control (women: 3,512 days, 121 people; men: 10,397 days, 951 people).

All analyses were conducted in Python (statsmodels).

## Data Availability

Data were provided by Juli Inc. under a data-sharing agreement and are not publicly available, owing to the sensitive nature of mental health and menstrual cycle data. Qualified researchers may request access from Juli Inc. for replication purposes, subject to a data use agreement. Requests should be directed to the corresponding author.

## Data availability

The data used in this study were provided by Juli Inc. under a data-sharing agreement and are not publicly available, owing to the sensitive nature of mental health and menstrual cycle data and to protect participant privacy. Access to the anonymised dataset may be requested from Juli Inc. by qualified researchers for the purpose of replicating the analyses reported here, subject to a data use agreement and applicable data protection requirements. Requests should be directed to the corresponding author.

## Code availability

The mcanalysis preprocessing and modelling package that is conceptually followed in this study is openly available (see reference [22]). Analysis scripts specific to this study are available from the corresponding author upon reasonable request.

## Acknowledgments

K.D.’s PhD studentship is funded by the Engineering and Physical Sciences Research Council (EPSRC). J.Z. is funded by the Pioneer Center for Statistical and Computational Methods for Advanced Research to Transform Biomedicine (SMARTbiomed). The funders played no role in the design of the study, the collection, analysis, or interpretation of data, or the writing of this manuscript.

## Author contributions

K.D. conceived and designed the study, conducted the statistical analysis, and wrote the manuscript. J.Z., J.H., A.K., N.K., and R.E. reviewed the manuscript and provided feedback on the analysis and interpretation, which K.D. incorporated in revisions. All authors read and approved the final manuscript.

